# Strategies used to manage overlap of primary study data by exercise-related overviews. Protocol for a systematic methodological review

**DOI:** 10.1101/2022.10.26.22281577

**Authors:** Ruvistay Gutierrez-Arias, Dawid Pieper, Carole Lunny, Rodrigo Torres-Castro, Raúl Aguilera-Eguía, Pamela Seron

## Abstract

**Introduction:** One of the most conflicting methodological issues when conducting an overview is the overlap of primary studies included across systematic reviews (SRs). Overlap in the pooled effect estimates across SRs may lead to overly precise effect estimates in the overview. SRs that focus on exercise-related interventions are often included in overviews aimed at grouping and determining the effectiveness of various interventions for the management of specific health conditions.

**Objective:** The aim of this systematic methodological review is to describe the strategies used by authors of overviews focusing on exercise-related interventions to manage the overlap of primary studies.

**Materials and methods:** A comprehensive search strategy has been developed for different databases and their platforms. The databases to be consulted will be MEDLINE (Ovid), Embase (Ovid), The Cochrane Database of Systematic Reviews (Cochrane Library), and Epistemonikos. Two reviewers will independently screen the records identified through the search strategy and will extract the information from the included overviews. The frequency and the type of overlap management strategies of the primary studies included in the SRs will be considered as the main outcome. In addition, the recognition of the lack of use of any overlap management strategy and the congruence between planning and conducting the overview focusing on overlap management strategies will be assessed. A sub-group analysis will be carried out using the impact factor of the journals at the time of publication of the overviews as the variable.

**Discussion:** This methodological review will provide a complete and comprehensive summary of the frequency of use and types of strategies used for managing the overlap of primary studies across the SRs included in the overviews focusing on exercise-related interventions in different health conditions. Future studies should apply different overlap management strategies to understand their impact on results and conclusions.

**Systematic review registration:** INPLASY202250161.

## Introduction

The number of published primary studies covering a similar research question has grown exponentially (1), limiting the possibility of keeping up to date on a specific topic (2). It is in this context that systematic reviews (SRs) with and without meta-analyses (MAs) of interventions can offer a solution (3), as in addition to synthesizing the available evidence, they use reproducible methods to assess the risk of bias in the primary studies included (4).

However, the number of published SRs and MAs has increased steadily in recent years despite the existence of repositories of SRs and MAs protocol registries (5–7) seeking to reduce duplication or redundancy of SR research (8,9).

The growth in research evidence makes it difficult for clinicians to stay current and use interventions based on the best available evidence (10,11). Overviews, also known as umbrella reviews, can help clinicians make sense of duplicated SRs on the same topic. Overviews synthesize information and data from multiple similar SRs to guide health decision-making (12).

Conducting overviews of health interventions is meant to map the available evidence (13), establishing the effects of different interventions on the same health condition or population (12), examining the effects of an intervention on different health conditions or populations (12), and determining the reasons for disagreement among SRs with or without MAs that answer the same research question (14).

Intuitively, one might think that conducting an overview presents the same steps as conducting an SR with MAs; however, overviews pose challenges stemming from the fact that the unit of analysis is the SR (15,16). When conducting an overview, one of the most conflicting methodological issues is the overlap of primary studies included across SRs with or without MAs (17). When one or more primary studies are included in two or more SRs with or without MAs, the results and conclusions of the overviews may be biased. Overlapping data from the same primary studies may include overlapping in risk of bias and certainty of evidence assessments (e.g., Grading of Recommendations, Assessment, Development and Evaluations (GRADE)), or overlapping in the determination of the effect of a specific intervention and other MA outcomes such as heterogeneity (e.g., I^2^) (18,19). Overlap in the pooled effect estimates across SRs may lead to overly precise effect estimates in the overview (20).

Methodological studies from different medical fields reported that authors of overviews rarely assess the overlap of primary studies (16,17). However, these studies have not conducted an exhaustive search of overviews oriented to a specific health problem, specialty, or discipline (16,17), as they have only searched an electronic database (16) and included heterogeneous overviews concerning the research questions addressed (16,17).

There are several ways to manage overlap (20). Some will depend heavily on the amount of overlap and the existing evidence base. Thus, it can be challenging to determine the methodological approach a priori. Changes to the protocol are likely to occur at this step and should be clearly reported.

SRs that focus on exercise-related interventions are often included in overviews aimed at grouping and determining the effectiveness of various interventions to mange of specific health conditions. Assessing the application of overlap management strategies in overviews focused on exercise-related interventions could contribute to identifying specific or differentiating aspects. This could be because the concept of exercise is often misunderstood (21). In addition, the existence of multiple interventions related to exercise due to their different modalities (e.g., continuous aerobic, intervallic aerobic, resistance exercise) and dosage (e.g., frequency, intensity, time, and type) could result in a particular need to manage the overlapping of primary studies data.

Therefore, this methodological review aims to find out how often strategies for handling overlapping data from primary studies are used across the systematic reviews considered by overviews focused on exercise-related interventions in different health conditions. Secondarily, we describe the overlap strategies used, the authors’ acknowledgment of not using any management strategy as a methodological weakness, and the congruence between the protocol and the final published overview in terms of overlap management.

## Materials and methods

The protocol of this methodological review is reported following the Preferred Reporting Items for Systematic Review and Meta-Analysis Protocols (PRISMA-P) (22) (see checklist in Supporting Information). In addition, this protocol has been registered in the International Platform of Registered Systematic Review and Meta-Analysis Protocols (INPLASY) under number INPLASY202250161.

### Eligibility criteria

Studies will be eligible if they meet the following inclusion criteria for study design and population. Given the purpose of this methodological review, the intervention and outcomes will not determine the inclusion of studies, and the comparator or control intervention will not be considered as it is not applicable.

#### Study design

We will include overviews that consider SRs with or without MAs, without distinction of the methodological design of the primary studies included. The definition of SR adopted by the authors of the overviews (23) will not be considered an eligibility criterion. Overviews that include primary studies not considered in the selected SRs will not be excluded.

For this review, an overview will be understood as any study (24) that:

1. synthesizes general information, methods, and outcome data from SRs, and
2. makes explicit the inclusion and exclusion criteria for SRs, and
3. includes an explicit search strategy for the studies, and
4. examines the effectiveness of health interventions.

Overviews that are conducted using a “rapid review” methodology (25) will be excluded, as the time frame in which they are conducted to answer urgent questions will likely not consider the overlap of the primary studies included in the SRs.

#### Population

Overviews include SRs that have considered primary studies that have studied any exercise-based intervention, where exercise is understood as a subcategory of physical activity that is planned, structured, repetitive, and purposefully focused on improving or maintaining one or more components of physical fitness (21), will be included. These overviews may include only SRs related to exercise-based interventions, or other non-exercise interventions as well.

Overviews that consider exercise training-based interventions that are applied both preventively and in the recovery phase, and that are delivered either as a stand-alone intervention, as part of a comprehensive rehabilitation program, or as an adjunct to other medical interventions in which exercise is the main component, will be included.

Furthermore, the inclusion of overviews will not be limited to the context in which the exercise-based interventions were applied (e.g., primary care, specialized care) or whether they were delivered face-to-face, remotely, or mixed.

Overviews that include SRs that consider physical activity as an intervention, understood as “any bodily movement produced by skeletal muscles that require energy expenditure” according to the World Health Organization (26), will be excluded. Therefore, to differentiate between exercise-based and physical activity-based interventions, it will be considered that the exercise, together with its structure and dosage (frequency, intensity, time, and type), must be prescribed or delivered by a professional related to physical training/rehabilitation.

#### Intervention

Our goal is to identify the strategies used to manage data from overlapping primary studies selected by SRs included in overviews. Strategies should be specified in the main text of the overviews and may be in the methods or results section, taking all possible methodological strategies that address overlap in the primary study data into consideration. Strategies addressing overlap can address different objectives (20), such as quantifying the overlap (17,27) (e.g., corrected covered area (CCA)), visually presenting overlap (28) (e.g., matrix, Venn and Euler diagrams), and avoiding duplicate information by using one or more decision algorithms (29) (e.g., quality of SRs, comprehensive SRs, up-to-datedness of SRs, statistical methods).

#### Outcomes

The presence and the type of overlap management strategies of the primary studies included in the SRs will be considered as the main outcome.

In addition, two aspects will be regarded as secondary outcomes:

1. Acknowledgement of the limitation in the conducting of the overview: we will assess whether the overview’s authors that did not include any strategy for managing primary study overlap considered this limitation in their discussion or conclusion.
2. Congruence between planning and conducting the overview: we will review available registry entries (e.g., PROSPERO) or published protocols in scientific journals (e.g., BMC Systematic Reviews Journal, BMJ Open) of all overviews included in this SR to determine whether management of primary study overlap had been considered in the planning phase of the overviews and to determine the congruence between the methods proposed in the protocols and those ultimately used.

### Search strategy

A search strategy translated to different databases and their platforms will be developed using a controlled vocabulary (MeSH and Emtree) and text words. The search strategy will include a search filter published in 2016 by Lunny et al. (30), which is validated to identify overviews in MEDLINE-Ovid with 93% sensitivity (95% CI 87 to 96). The search strategy constructed for this database and platform is shown in Table N°1, which will be used as a basis for adapting the search strategies of the other databases and search platforms.

**Table 1.**
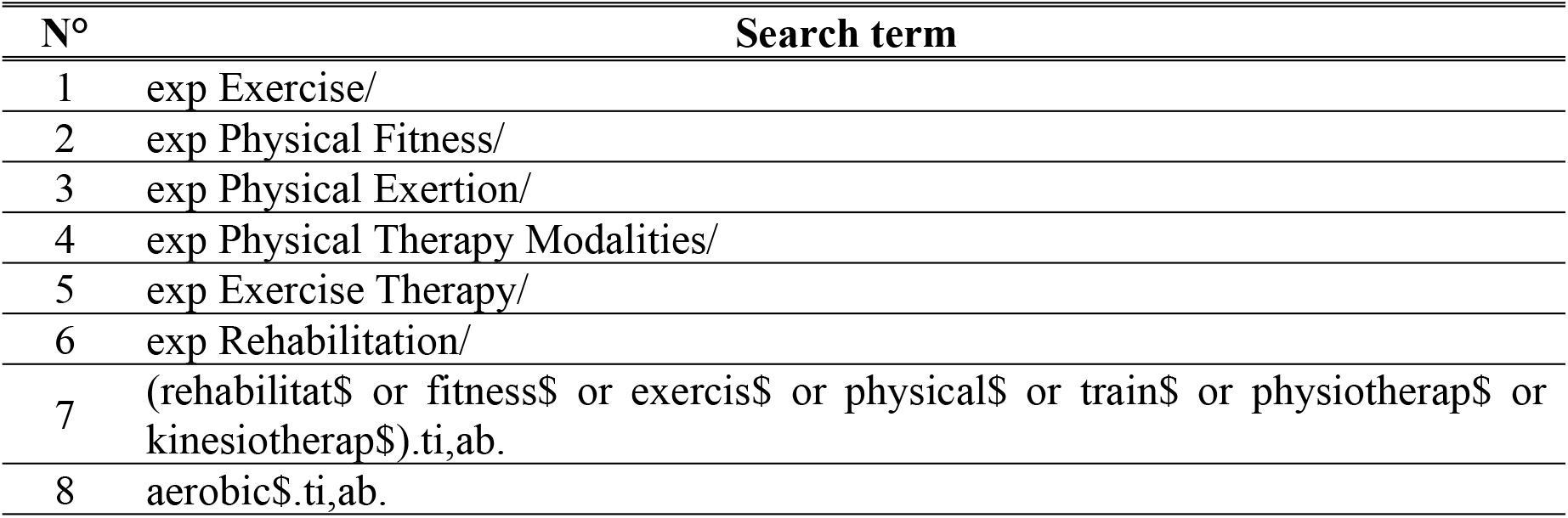
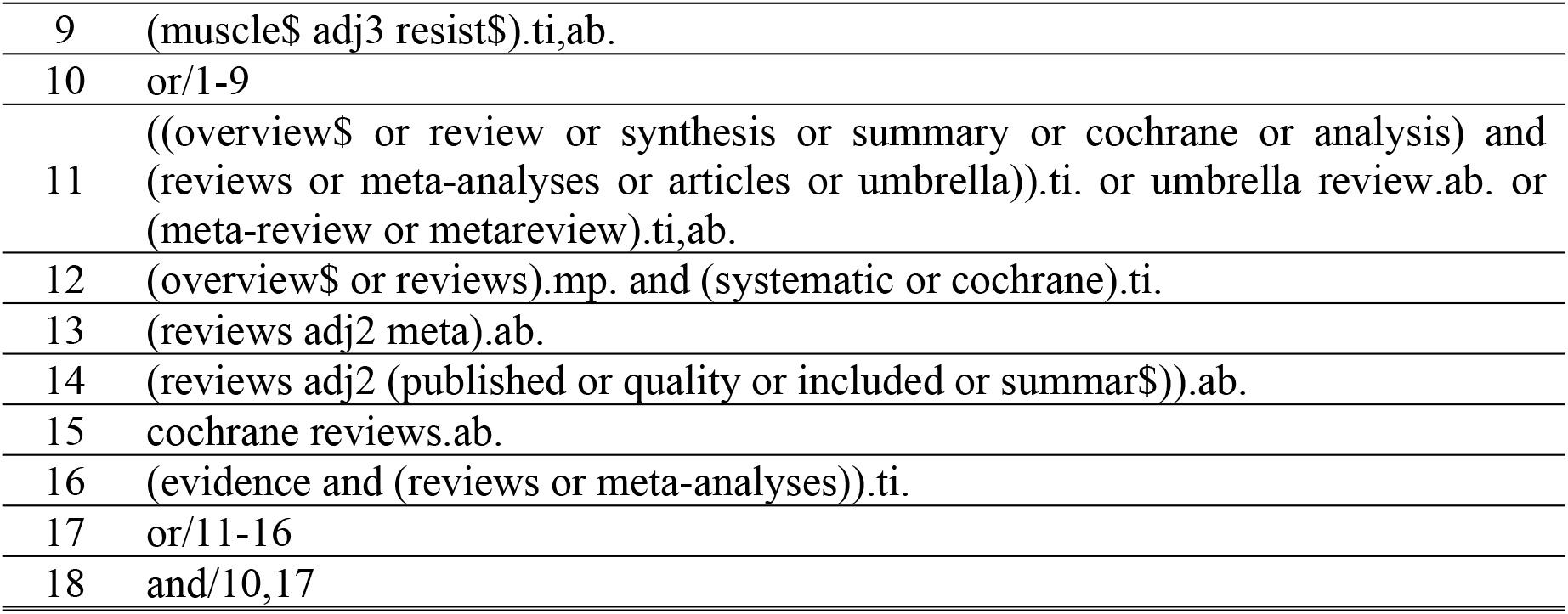
Search strategy for MEDLINE using the Ovid platform

| N° | Search term |
| --- | --- |
| 1 | exp Exercise/ |
| 2 | exp Physical Fitness/ |
| 3 | exp Physical Exertion/ |
| 4 | exp Physical Therapy Modalities/ |
| 5 | exp Exercise Therapy/ |
| 6 | exp Rehabilitation/ |
| 7 | (rehabilitat\$ or fitness\$ or exercis\$ or physical\$ or train\$ or physiotherap\$ or kinesiotherap\$).ti,ab. |
| 8 | aerobic\$.ti,ab. |
| 9 | (muscle\$ adj3 resist\$).ti,ab. |
| 10 | or/1-9 |
| 11 | ((overview\$ or review or synthesis or summary or cochrane or analysis) and (reviews or meta-analyses or articles or umbrella)).ti. or umbrella review.ab. or (meta-review or metareview).ti,ab. |
| 12 | (overview\$ or reviews).mp. and (systematic or cochrane).ti. |
| 13 | (reviews adj2 meta).ab. |
| 14 | (reviews adj2 (published or quality or included or summar\$)).ab. |
| 15 | cochrane reviews.ab. |
| 16 | (evidence and (reviews or meta-analyses)).ti. |
| 17 | or/11-16 |
| 18 | and/10,17 |

The databases to be consulted will be MEDLINE (Ovid), Embase (Ovid), The Cochrane Database of Systematic Reviews (Cochrane Library), and Epistemonikos. In addition, we will search protocol registries of SRs such as the International Platform of Registered Systematic Review and Meta-analysis Protocols (INPLASY) (https://inplasy.com/), PROSPERO (https://www.crd.york.ac.uk/PROSPERO/), and OSF Registries (https://osf.io/registries), and follow up protocols published in scientific journals (e.g., BMC Systematic Reviews Journal, BMJ Open).

We will also review the references of the studies included in this review to identify overviews that may not have been identified by our electronic search strategy.

We will include all languages in our search and will not be limited by the date of publication/indexing in databases.

### Study selection

Two reviewers (RGA and RTC) will independently and blindly screen the records identified through the search strategy. In the first instance, the titles and abstracts will be evaluated for inclusion. Then the full texts of the records qualified as potentially eligible, and those that did not present sufficient information to be excluded, will be checked for compliance with all eligibility criteria. The Rayyan® application (31) will be used for this stage. Disagreements will be resolved by consensus, or ultimately by a third-party reviewer (RAE or PS).

### Data extraction

The extraction of information from the included overviews will also be carried out independently and blindly by two reviewers (RGA and RTC). For this, a standardized extraction form will be used which will contain data related to the basic information of the overviews:

- Title.
- Journal name.
- Year of publication.
- Name of the authors.
- Objectives of SRs.
- Number of SRs included
- Methodological aspects: databases consulted, date of search, type of synthesis of results (narrative, MA, or both), and instruments for assessing the risk of bias/methodological quality of the SRs included.

Data will be extracted to respond to the findings of this methodological review:

- Type of overlap management strategy:
  a. Quantifying overlap: e.g., CCA.
  b. Visual presentation of the overlap: e.g., matrix, Venn and Euler diagrams.
  c. Strategies to avoid duplicate information: e.g., Algorithms based on the quality of SRs, comprehensive SRs, up-to-datedness of SRs, statistical methods such as sensitivity analyses, or a combination of two or more criteria.
- Step in the conducting of the overview where the strategy has been deployed or used: e.g., data extraction step, synthesis step.
- Level at which the strategies were applied: i.e., whether it was at the level of SR or reported outcomes (20).

In addition, the impact factor (IF) of the journal at the time of publication of the overviews will be recorded. This will be extracted from the journals’ official websites or from Web of Science (https://www.webofscience.com/).

If more than one record or publication exists for an overview, the most recent version will be considered for analysis. Disagreements will be resolved by consensus, or ultimately by a third-party reviewer (RAE or PS).

### Risk of bias assessment

This methodological review assesses one aspect that may affect the methodological quality or risk of bias of the overviews. The assessment of the overall risk of bias of the overviews is not an objective of this study.

### Strategy for data synthesis

The results of the study selection will be schematized through a PRISMA-type flow chart (32). In addition, the characteristics of the overviews included, as well as data related to the primary and secondary outcomes, will be presented in narrative form, and through tables and figures.

Descriptive statistics will be used to quantify the number of overviews using overlap strategies, whether the strategies were used at the level of the SRs or the level of each reported outcome. In addition, these results will be organized by the type of strategy used.

We will also assess whether the overlapping strategy successfully resolved overlap at the following steps: risk of bias assessment, the certainty of the evidence (e.g., GRADE), and the synthesis step.

### Analysis of subgroups

Differences in the percentage of overviews that include overlap management strategies, the type of strategies used, the recognition of the weakness of not using any strategy, and the congruence between the protocols and the methodology finally used among journals with and without IF will be assessed. In addition, this analysis will be repeated for IF journals, considering the median or quartiles of the IF of the journals at the time of publication of the overviews to form 2 or 4 groups respectively, depending on the number of overviews included in this methodological review.

## Discussion

This methodological review will provide a comprehensive and exhaustive summary of the frequency of use of strategies for managing primary study overlap across SRs included in overviews focused on exercise-related interventions in different health conditions. It will also provide insight into the strategies used to quantify and visualize overlap, as well as those used to avoid duplicate data.

On the other hand, the findings of this review will tell us whether the authors of the overviews recognized the failure to include some strategy for handling overlap as a methodological weakness, taking into account that the greater the degree of overlap, the more falsely precise the estimates of the effects of the interventions (20). In addition, the congruence between the strategies used by the published overviews and their respective protocols will be revealed. To our knowledge, the latter two aspects have not been addressed at the overview level by other studies before.

### Future research

Different overlapping data management strategies will be applied to all, or a representative sample, of the overviews identified by this methodological review.

In addition, it would be interesting for future studies to assess the association between the use of different strategies for handling overlapping primary studies and methodological quality of the overviews or compliance with recommendations in overview reporting, such as the PRIOR statement (33).

### Dissemination plans

The findings of this review will be presented at scientific conferences and published as one or more studies in peer-review scientific journals related to rehabilitation, healthcare, or methodological aspects associated with evidence synthesis.

## Data Availability

No datasets were generated or analysed during the current study. All relevant data from this study will be made available upon study completion.

## Acknowledgments

None

## Notes

### Competing Interest Statement

The authors have declared no competing interest.

### Funding Statement

The author(s) received no specific funding for this work.

### Author Declarations

This protocol does not require an ethics statement.

